# Performance of Advanced Large Language Models (GPT-4o, GPT-4, Gemini 1.5 Pro, Claude 3 Opus) on Japanese Medical Licensing Examination: A Comparative Study

**DOI:** 10.1101/2024.07.09.24310129

**Authors:** Mingxin Liu, Tsuyoshi Okuhara, Zhehao Dai, Wenbo Huang, Hiroko Okada, Emi Furukawa, Takahiro Kiuchi

## Abstract

**Purpose:** This study aims to evaluate the accuracy of medical knowledge in the most advanced LLMs (GPT-4o, GPT-4, Gemini 1.5 Pro, and Claude 3 Opus) as of 2024. It is the first to evaluate these LLMs using a non-English medical licensing exam. The insights from this study will guide educators, policymakers, and technical experts in the effective use of AI in medical education and clinical diagnosis.

**Method:** Authors inputted 790 questions from Japanese National Medical Examination into the chat windows of the LLMs to obtain responses. Two authors independently assessed the correctness. Authors analyzed the overall accuracy rates of the LLMs and compared their performance on image and non-image questions, questions of varying difficulty levels, general and clinical questions, and questions from different medical specialties. Additionally, authors examined the correlation between the number of publications and LLMs’ performance in different medical specialties.

**Results:** GPT-4o achieved highest accuracy rate of 89.2% and outperformed the other LLMs in overall performance and each specific category. All four LLMs performed better on non-image questions than image questions, with a 10% accuracy gap. They also performed better on easy questions compared to normal and difficult ones. GPT-4o achieved a 95.0% accuracy rate on easy questions, marking it as an effective knowledge source for medical education. Four LLMs performed worst on “Gastroenterology and Hepatology” specialty. There was a positive correlation between the number of publications and LLM performance in different specialties.

**Conclusions:** GPT-4o achieved an overall accuracy rate close to 90%, with 95.0% on easy questions, significantly outperforming the other LLMs. This indicates GPT-4o’s potential as a knowledge source for easy questions. Image-based questions and question difficulty significantly impact LLM accuracy. “Gastroenterology and Hepatology” is the specialty with the lowest performance. The LLMs’ performance across medical specialties correlates positively with the number of related publications.

## Introduction

Since the release of the first artificial intelligence (AI) chatbot based on a large language model (LLM), ChatGPT, by OpenAI in November 2022, various LLMs, such as GPT-3.5, GPT-4, Gemini, Claude, and Llama, have swiftly garnered global attention based on their ability to provide detailed answers to complex queries ^1–4^. These LLMs have found widespread applications in programming, law, education, and business, demonstrating impressive performance ^5–8^. In the medical field, LLMs have shown potential in clinical diagnosis and medical education. For instance, compared with traditional search engines, which present a list of relevant pages, LLM-based AI chatbots aim to provide concise and practical answers to users’ questions, making them effective knowledge resources ^9, 10^. Additionally, most advanced LLMs possess image analysis capabilities, suggesting their potential as tools for diagnosing and analyzing medical images, such as skin rashes and X-ray images ^9^. However, the accuracy of the medical knowledge possessed by large-language artificial intelligence models represents the final barrier to their practical application in medical education and clinical diagnosis. A previous study indicated that for LLMs to be used as effective knowledge resources in medical education, the accuracy of the responses must consistently exceed 95% ^11^. Researchers have used medical licensing examination questions from various countries to assess the accuracy of medical knowledge possessed by LLMs ^12–21^. Among these studies, the most frequently tested LLMs were GPT-4 and GPT-3.5. A systematic review showed that GPT-4 can achieve an overall accuracy rate of 81% across medical licensing exams from different countries, successfully passing most of these exams ^22^. However, it falls significantly below the 95% accuracy threshold. For most image-based questions, GPT-4’s accuracy is even below 70% ^22^. More than a year has passed since GPT-4’s release on March 14, 2023. By 2024, more advanced LLMs have been available to the public. Anthropic released the Claude 3 Opus on March 4 ^23^.

OpenAI and Google released GPT-4o and Gemini 1.5 Pro on May 13, 2024, and May 14, 2024, respectively. According to the MMLU benchmark results, Claude 3 Opus scored 86.8% and GPT-4o scored 88.7%, both surpassing GPT-4’s score of 86.4% ^23–25^. Gemini 1.5 Pro scored 85.9%, which is similar to GPT-4 ^2^. We hypothesized that more advanced LLMs would achieve higher accuracy rates in medical licensing examinations, approaching or surpassing the 95% threshold. A previous study used the United States Medical Licensing Examination (USMLE) to test GPT-4o’s accuracy in answering medical licensing exam questions in English. GPT-4o scored between 85.7% and 92.5% across the three steps of the USMLE ^26^. However, the performance of the GPT-4o on medical licensing exams in English-speaking countries may not fully reflect its performance on medical licensing exams in non-English-speaking countries.

Furthermore, this study did not test other LLMs with performance similar to that of GPT-4o. To the best of our knowledge, no study has tested the performance of the most advanced LLMs on medical licensing exams in non-English-speaking countries in 2024. Our study aimed to fill this gap in the literature.

### Study Aims and Objectives

Our study utilized the Japanese Medical Licensing Examination (JNME) to evaluate the performance of the four most advanced LLMs, GPT-4o, Claude 3 Opus, Gemini 1.5 Pro and GPT-4, to clarify the following issues:

1. How do GPT-4o, Claude 3 Opus, and Gemini 1.5 Pro compare with GPT-4 in terms of overall performance? Which LLM method performs best? Can any LLM pass the exam or reach a 95% accuracy threshold?
2. How do LLMs perform on image-based versus non-image-based questions?
3. Does the year of publication of the questions affect LLM performance?
4. How do LLMs perform in different medical specialties?
5. Does question difficulty influence LLM performance?
6. Does the number of publications in various medical specialties correlate with LLM performance in these specialties?

By addressing these questions, we will deepen our understanding of LLMs in answering medical questions, summarizing existing issues, and providing recommendations for the future development and updating of LLMs. We aim to pave the way for the application of LLMs in medical education and clinical diagnosis.

## Methods

### Tested LLMs

We selected the four most advanced LLMs as of June 2024—GPT-4o, GPT-4, Gemini 1.5 Pro, and Claude 3 Opus—as test subjects ^2, 23, 24^.

### Japanese National Medical Examination (JNME)

The JNME was initiated in 1946 as an examination for medical school graduates to obtain a national medical license to become doctors after six years of advanced training. The JNME format has undergone several changes in recent years. In 2018, it adopted its current format. The latest version of the JNME comprises 400 questions divided into six sections labeled A–F. Sections B and E contain essential questions, each comprising 50 questions. Sections A, C, D, and F contain nonessential questions, each comprising 75 questions. The exam questions are also categorized into general and clinical questions. In the essential sections (B and E), general questions are worth one point each, while clinical questions are worth three points each, with a passing score of 160 points. In the nonessential sections (A, C, D, and F), all questions are worth one point, and the passing score is not fixed and is usually around 220 points. Additionally, the exam includes taboo questions, in which making more than three mistakes results in failure. The annual pass rate for Japanese medical students is approximately 90%. The question types include multiple-choice questions (MCQs) and calculation questions. MCQs are categorized into five options with a single-answer, five options with two- or three answers (select two or three), and MCQs with more than six options. For multiple-answer questions, the prompt indicates the number of correct choices. Image-based questions are also included in the examination ^27^.

### Questions Used in this Study

To clarify whether the cut-off date of LLMs affects their performance, this study used the earliest exam (JNME 2018) and the latest exam (JNME 2024) in the latest format. Because the cutoff dates for the training data of GPT-4o, GPT-4, Gemini 1.5 Pro, and Claude 3 Opus are October 2023, April 2023, November 2023, and August 2023, respectively, the questions and answers for JNME 2024 held on February 7, 2024, are unknown to these large language models ^28–30^. By contrast, JNME 2018 may be part of the training data for these models. We can compare the performances of large language models in exams conducted before and after the cutoff dates of their training data. Questions that were deemed incorrect and image-based questions with nonpublic images were excluded. We categorized the exam questions according to the following features:

1. Image-based and non-image-based questions.
2. General and clinical questions.
3. A co-author with a Japanese medical license (ZH Dai) classified all questions into 21 medical specialties based on Web of Science categories ^31^.
4. According to the correct answer rate among the human examinees of each exam question published by Medu4 ^32^, a preparatory school for the JNME, questions were categorized into three difficulty levels: easy (accuracy rate ≥ 90%), medium (accuracy rate ≥ 70% and < 90%), and difficult (accuracy rate < 70%).

### Inputting Questions to LLMs

We inputted Questions into the LLMs from June 2 to 16, 2024. Question texts and images were directly inserted into the chat window of each LLM, rather than using an Application Programming Interface (API). The questions were given in the same order as in the actual exam. Each question was asked only once. In cases where the LLMs were unable to provide an answer for system reasons, we re-asked the question until the LLMS provided a response. To avoid the context of previous chats from affecting the accuracy of answers, each question was asked in a new independent chat. However, questions of continuity were asked within the same chat. All questions were asked in the original Japanese language. No prompts were used when imputing questions to GPT-4o, GPT-4, and Claude 3 Opus, no prompt was used. As 23 questions involved two or more images, and Gemini 1.5 Pro could upload only one image at a time, we used the prompt “Please review two images and answer the related question. This is the first/second image.” Additionally, when the LLMs refused to answer a question, we used the prompt “This is a question from the medical licensing examination” to obtain a response. All answers were collected in an Excel spreadsheet and the answers to each question were marked as correct or incorrect by two authors (MX Liu and WB Huang).

### Number of Publications Across Each Medical Specialty

We used the Web of Science (WOS) category features within the WOS Core Collection, one of the largest document repository databases, to retrieve the number of publications in 21 medical specialties ^33^. The search covered publications from January 1, 1900, to December 31, 2023. We recorded the total number of publications, articles, and open-access articles in each medical specialty.

### Statistical Analysis

Descriptive statistics, including the number of questions, correct answers, proportion, mean value, and 95% confidence interval (CI) were employed to represent the performance of each LLM. The accuracy rates from different LLMs and categories were compared using Fisher’s exact test. Pearson’s correlation coefficient was used to detect the association between LLMs and the number of publications. Significance was defined as P<0.05 using two-tailed tests. All statistical analyses were performed using R version 4.4.0.

### Ethical Considerations

The JNME questions and LLMs used in this study are publicly accessible online. Therefore, ethical approval was not required.

## Results

### Characteristic of JNME Questions

In the JNME 2018, two questions deemed incorrect and eight questions with non-public images were excluded, leaving 390 questions included in the test. All 400 questions from the 2024 examination were included in the test. Of the 790 questions, 293 were general and 497 were clinical. In total, 199 image-based questions were asked. The numbers of easy, normal, and difficult questions were 433, 217, and 130, respectively. The specific number of questions for each category is presented in Supplementary Materials 1.

### The Accuracy Rate of LLMs’ Response

GPT-4o, GPT-4, and Claude 3 Opus answered all 790 questions without prompt. Gemini 1.5 Pro refused to answer the 12 questions that included images of human body parts. In these cases, prompts were used to obtain responses. However, Gemini 1.5 Pro refused to answer two questions even with prompts. Therefore, the Gemini 1.5 Pro provided answers to 788 questions. All responses and their correct markings are presented in Supplementary Materials 2 and 3.

The overall accuracy rates for the four LLMs, from highest to lowest, were 89.2% (95% CI: 86.90%-91.30%) for GPT-4o, 82.0% (95% CI: 79.20%-84.60%) for Claude 3 Opus, 80.1% (95% CI: 73.70%-82.20%) for Gemini 1.5 Pro, and 76.8% (95% CI: 73.70%-79.70%) for GPT-4. The accuracy rate of GPT-4o was significantly higher than that of the other three LLMs (all P-values < 0.001), and it achieved an accuracy rate of over 90.0% (90.8%) for the 2024 JNME. All four LLMs had higher accuracy rates on the 2024 JNME compared to the 2018 JNME, but the differences were not statistically significant (all P-values ≥ 0.05). (Table 1)

**Table 1.** Overall accuracy rate of LLMs.

| **GPT-4o** | **GPT-4** | **Gemini 1.5 PRO** | **Claude 3 Opus** |
| --- | --- | --- | --- |

|  | Correct<br>number | Correct<br>rate,<br>95%CI | Correct<br>number | Correct<br>rate,<br>95%CI | Correct<br>number | Correct<br>rate,<br>95%CI | Correct<br>number | Correct<br>rate,<br>95%CI |
| --- | --- | --- | --- | --- | --- | --- | --- | --- |
| **JNME** |  | 90.8% |  | 77.5% |  | 82% |  | 82% |
| **2024** | 363 | (87.50%- | 310 | (73.10%- | 327/399 | (77.80%- | 328 | (77.90%- |
| **(n=400)** |  | 93.40%) |  | 81.50%) |  | 85.60%) |  | 85.60%) |
| **JNME** |  | 87.7% |  | 76.2% |  | 78.1% |  | 82.1% |
| **2018** | 342 | (84.00%- | 297 | (71.60%- | 304 | (73.70%- | 320 | (77.90%- |
| **(n=390)** |  | 90.80%) |  | 80.30%) |  | 82.20%) |  | 85.70%) |
| **JNME** |  |  |  |  |  |  |  |  |
| **2024** |  | 89.2% |  | 76.8% |  | 80.1% |  | 82% |
| **and** | 705 | (86.90%- | 607 | (73.70%- | 631 | (73.70%- | 648 | (79.20%- |
| **2018** |  | 91.30%) |  | 79.70%) |  | 82.20%) |  | 84.60%) |
| **(n=790)** |  |  |  |  |  |  |  |  |

We calculated the scores of the four LLMs based on the scoring rules of JNME and compared them with the passing threshold. GPT-4O, Gemini 1.5 Pro, and Claude 3 Opus passed the 2024 JNME exam, whereas GPT-4 missed the passing score by one point on the nonessential questions. Claude 3 Opus achieved a passing score on the nonessential questions of the 2024 JNME. All four LLMs passed the 2018 exam, with GPT-4 exceeding the passing score by only two points on the nonessential questions. (Table 2)

**Table 2.** Score of each LLM calculated by JNME scoring rules.

|  | Category | Pass line<br>of the<br>exam | GPT-4o | GPT-4 | Gemini 1.5<br>PRO | Claude 3<br>Opus |
| --- | --- | --- | --- | --- | --- | --- |
| **JNME**<br>**2024**<br>**(n=400)** | Essential questions | 160 | 185 | 167 | 182 | 179 |
|  | Nonessential questions | 230 | 270 | 229 | 238 | 230 |
| **JNME**<br>**2018**<br>**(n=390)** | Essential questions | 160 | 193 | 174 | 174 | 176 |
|  | Nonessential questions | 208 | 246 | 210 | 217 | 231 |

Among all 790 questions, the accuracy rates for non-image-based questions were as follows: 92.2% (95% CI: 89.8%-94.2%) for GPT-4o, 80.0% (95% CI: 76.6%-83.2%) for GPT-4, 81.9% (95% CI: 78.5%-84.9%) for Gemini 1.5 Pro, and 83.6% (95% CI: 80.4%-86.5%) for Claude 3 Opus. All values were above 80%, with GPT-4o achieving an accuracy of over 90%. For image-based questions, the accuracy rates were 80.4% (95% CI: 74.2%-85.7%) for GPT-4o, 67.3% (95% CI: 60.4%-73.8%) for GPT-4, 74.6% (95% CI: 67.9%-80.5%) for Gemini 1.5 Pro, and 77.4% (95% CI: 70.9%-83.0%) for Claude 3 Opus. Only GPT-4o had an accuracy rate above 80% for image-based questions. The accuracy rates for the non-image-based questions were higher by 11.8%, 12.7%, 7.4%, and 6.2% for the GPT-4o, GPT-4, Gemini 1.5 Pro, and Claude 3 Opus, respectively. Three of them showed significant differences (GPT-4O and GPT-4: P<0.001; Gemini 1.5 Pro: P=0.03; Claude 3 Opus: P=0.054). By separately examining the results for 2018 and 2024, all four LLMs had higher accuracy rates for non-image-based questions than image-based questions. (Table 3)

**Table 3.**
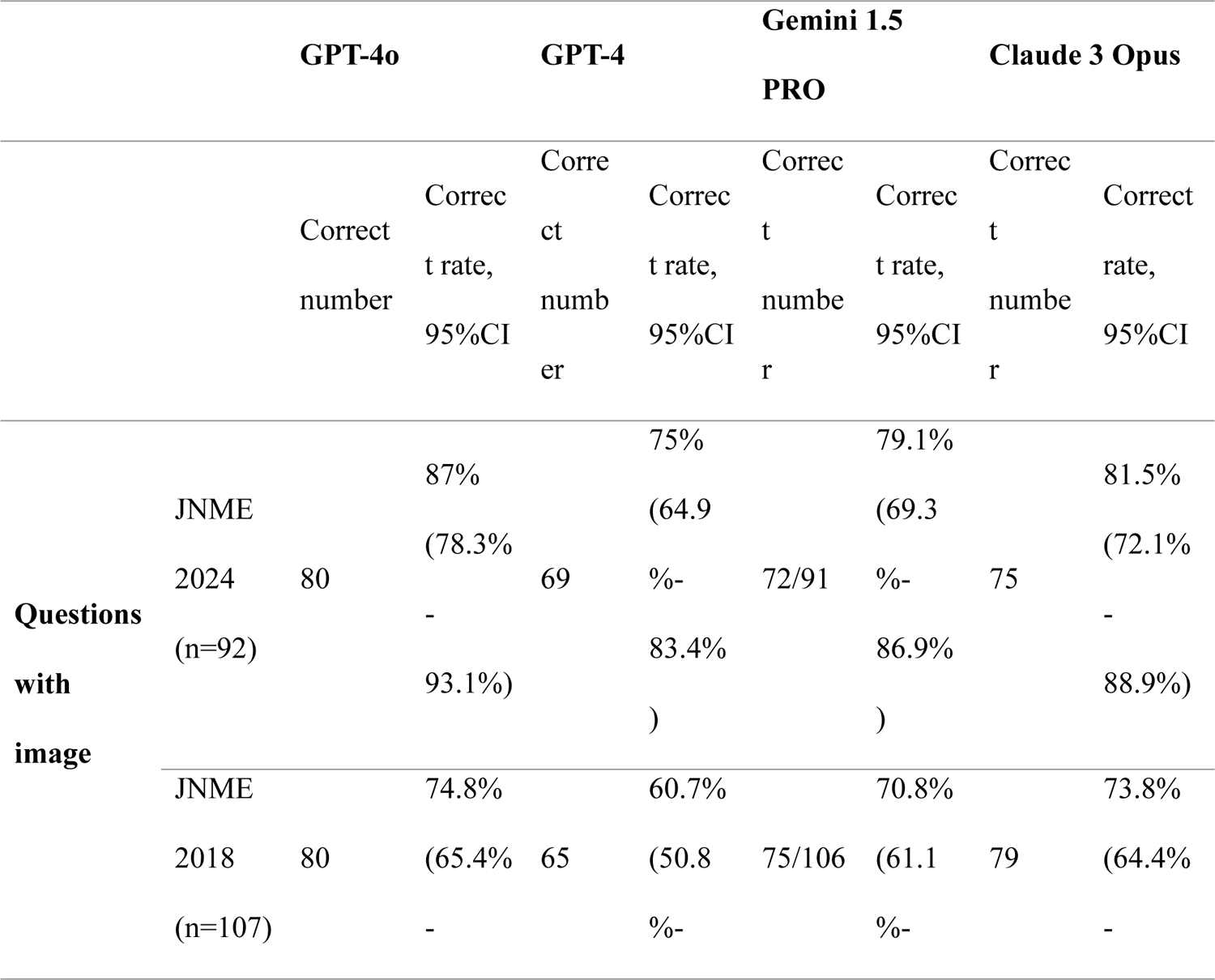
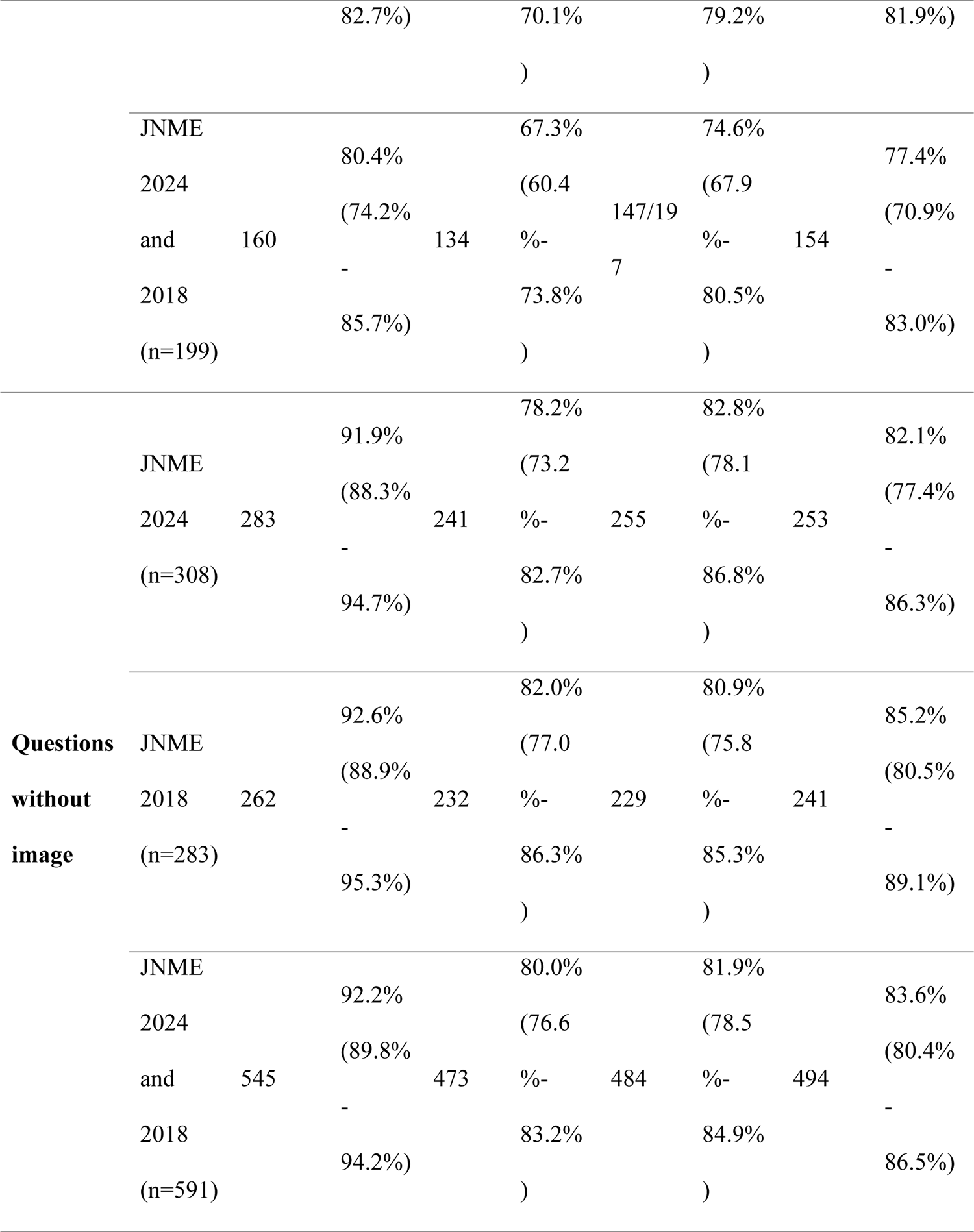
Accuracy rate of each LLM on non-image-based questions and image-based questions.

For performance on questions with different difficulty levels, GPT-4o’s accuracy rates were as follows: 95.0% (95% CI: 92.6%–96.9%) for easy questions, 85.3% (95% CI: 79.8%–89.7%) for normal questions, and 76.2% (95% CI: 67.0%–83.2%) for difficult questions. GPT-4’s accuracy rates were: 84.2% (95% CI: 80.5%–87.5%) for easy questions, 71.0% (95% CI: 64.4%–76.9%) for normal questions, and 61.5% (95% CI: 52.6%–69.9%) for difficult questions. The accuracy rates of Gemini 1.5 Pro were 86.8% (95% CI: 82.9%–89.5%) for easy questions, 77.9% (95% CI: 71.8%–83.2%) for normal questions, and 61.5% (95% CI: 52.6%–69.9%) for difficult questions. Claude 3 Opus accuracy rates were 89.2% (95% CI: 85.9%–91.9%) for easy questions, 76.0% (95% CI: 69.8%–81.6%) for normal questions, and 67.7% (95% CI: 58.9%– 75.6%) for difficult questions. For all four LLMs, the accuracy rate differences between easy and normal questions and between normal and difficult questions were approximately 10%, showing statistically significant differences (all p< 0.05). (Table 4)

**Table 4.**
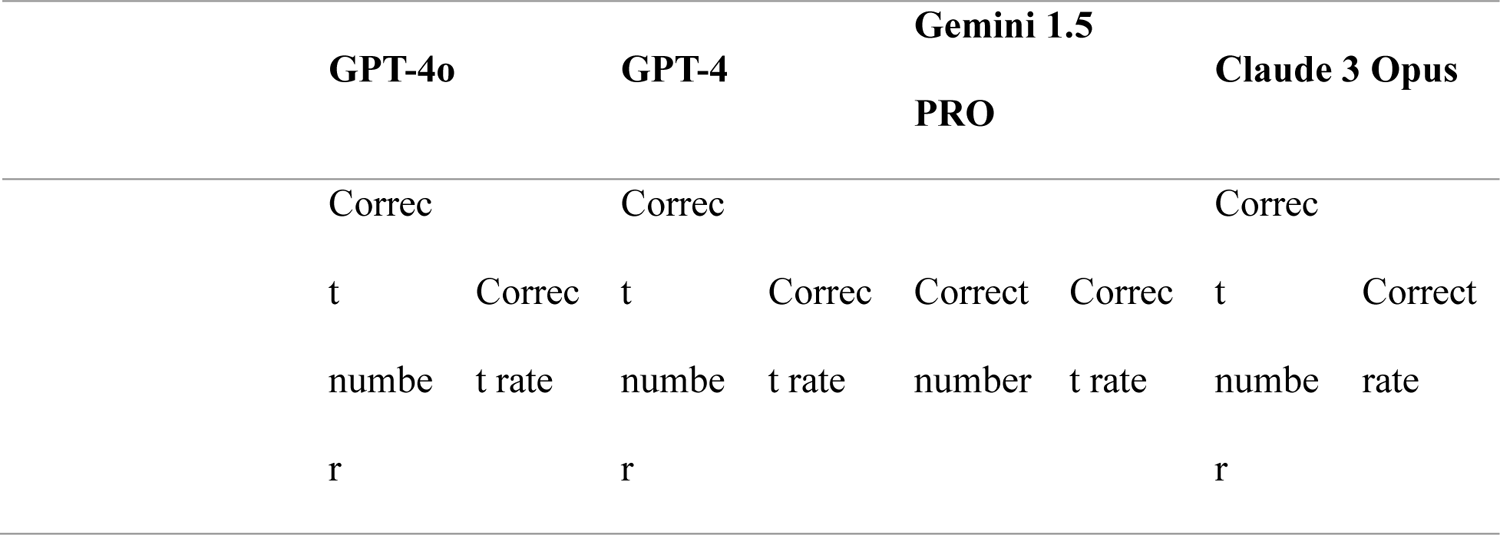
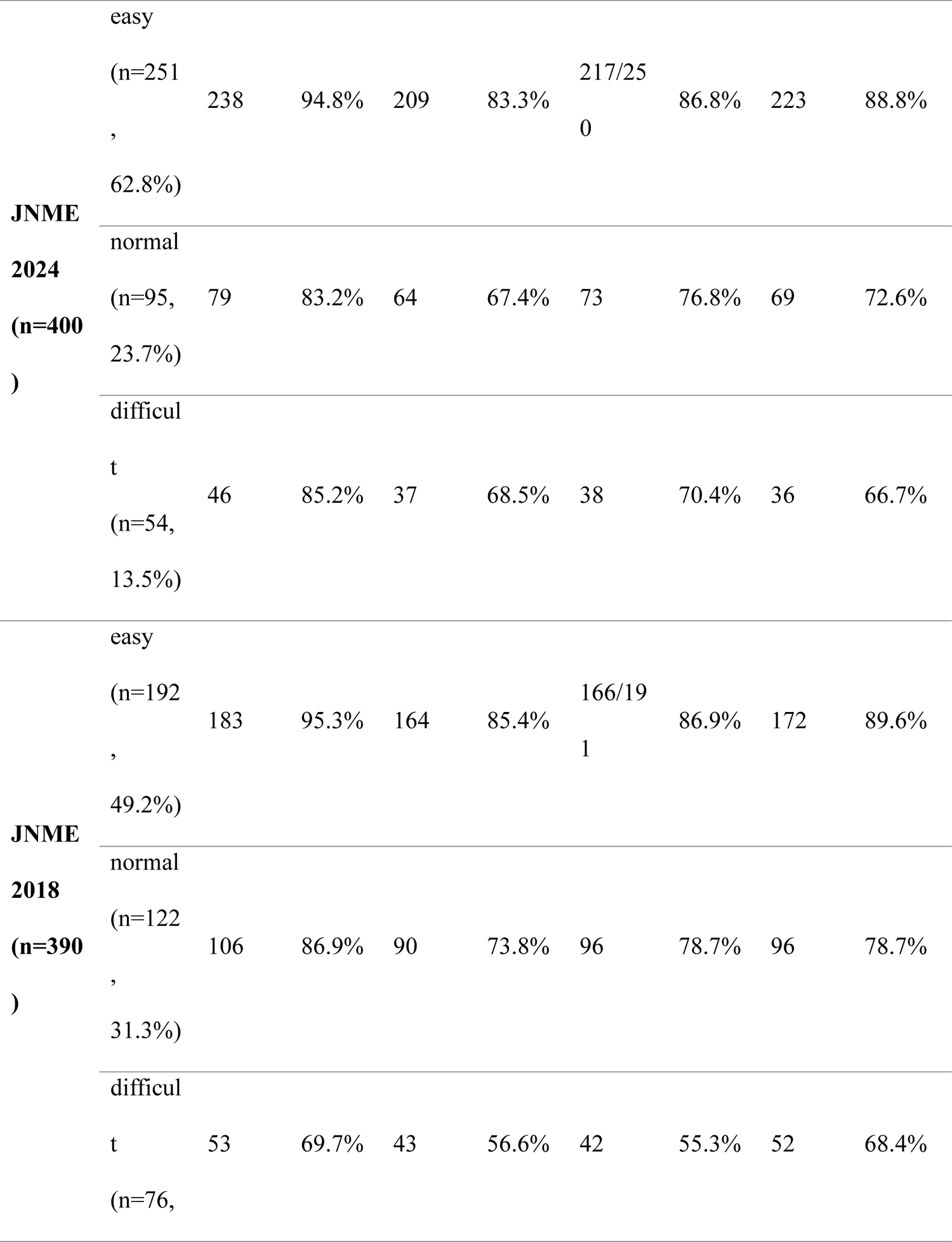
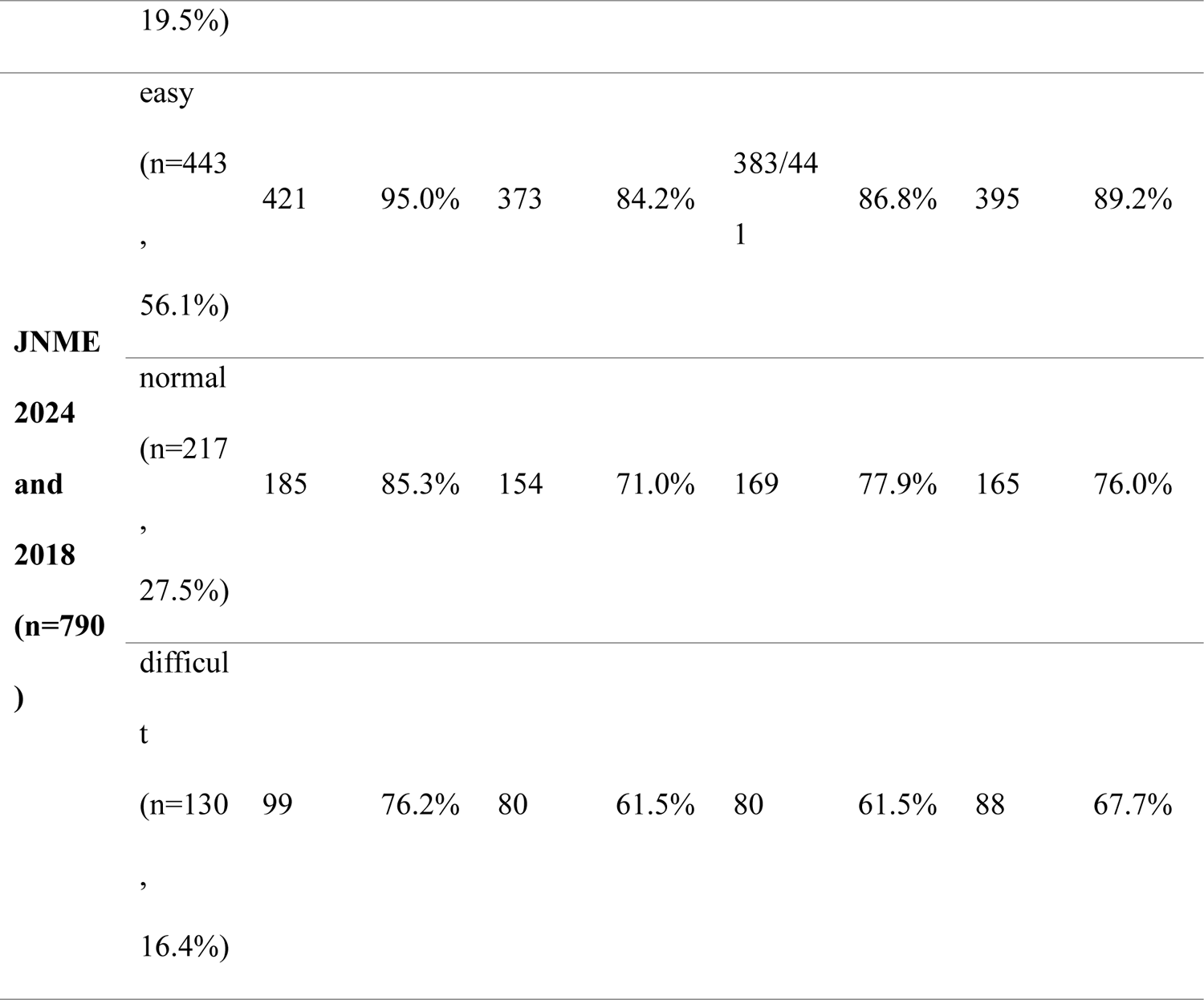
Accuracy rate of each LLM on different difficulty level questions.

GPT-4o had an accuracy rate that was 3.6% higher for general questions than for clinical questions. GPT-4, Gemini 1.5 Pro, and Claude 3 Opus had accuracy rates for clinical questions that were 5.5%, 1.7%, and 2.9% higher than those for general questions, respectively. None of these differences was statistically significant (p > 0.05). (Table 5)

**Table 5.** Accuracy rate of each LLM on general and clinical questions.

| GPT-4o | GPT-4 | Gemini 1.5 PRO | Claude 3 Opus |
| --- | --- | --- | --- |

|  |  | Correct<br>number | Correct<br>rate | Correct<br>number | Correct<br>rate | Correct<br>number | Correct<br>rate | Correct<br>number | Correct<br>rate |
| --- | --- | --- | --- | --- | --- | --- | --- | --- | --- |
| **JNME**<br>**2024**<br>**(n=400)** | General |  |  |  |  |  |  |  |  |
|  | questions<br>(n=147) | 134 | 91.2% | 101 | 68.7% | 116/146 | 79.5% | 110 | 74.8% |
|  | Clinical |  |  |  |  |  |  |  |  |
|  | questions<br>(n=253) | 229 | 90.5% | 209 | 82.6% | 211 | 83.4% | 218 | 86.2% |
| **JNME**<br>**2018**<br>**(n=390)** | General |  |  |  |  |  |  |  |  |
|  | questions<br>(n=146) | 134 | 91.8% | 114 | 78.1% | 115/145 | 79.3% | 125 | 85.6% |
|  | Clinical |  |  |  |  |  |  |  |  |
|  | questions<br>(n=244) | 208 | 85.2% | 183 | 75.0% | 189 | 77.5% | 195 | 79.9% |
| **JNME**<br>**2024**<br>**and**<br>**2018**<br>**(n=790)** | General |  |  |  |  |  |  |  |  |
|  | questions<br>(n=293) | 268 | 91.5% | 215 | 73.4% | 231/291 | 78.8% | 235 | 80.2% |
|  | Clinical |  |  |  |  |  |  |  |  |
|  | questions<br>(n=497) | 437 | 87.9% | 392 | 78.9% | 400 | 80.5% | 413 | 83.1% |

The medical specialties in which the accuracy of all four LLMs was higher than their overall average accuracy, ranked by average accuracy from highest to lowest, are as follows: Emergency medicine (including toxicology), Anesthesiology, and Intensive Care (91.5%), Cardiology (91.4%), Immunology and Rheumatology (90.7%), Ophthalmology (89.7%), Orthopedics (88.6%), Infectious disease (80.6%), Others or general knowledge (85.0%), and Psychiatry (84.8%). Conversely, the medical specialties in which the accuracy of all four LLMs was lower than their overall average accuracy, ranked by average accuracy from lowest to highest, are: Gastroenterology and Hepatology (65.2%), Hematology (71.4%), Endocrinology and Metabolism (75.0%), Pulmonology (75.6%), Dermatology (77.1%), and Obstetrics and Gynecology (78.1%). Among these, the accuracy rate of Gastroenterology and Hepatology (P<0.001), Cardiology (P<0.001), Emergency medicine (including toxicology), Anesthesiology and Intensive Care (P<0.01), Immunology and Rheumatology (P=0.03), Pulmonology (P=0.046), and Hematology (P=0.02) show significant differences compared to the overall average accuracy (82.0%). (Supplementary Materials 4 and Figure 1).

### Correlation Between Accuracy and the Number of Publications of Each Medical Specialty

LLMs’ accuracy across various medical specialties was positively correlated with the number of all types of literature, articles, and open-access articles. Among the three types of literature, the correlation between the four LLMs and number of articles was the highest, followed by open-access articles, with the correlation with all types of literature being the lowest. Only Gemini 1.5 Pro showed a significant positive correlation between accuracy in each medical specialty and the number of articles and open-access articles (P=0.03). The correlations with the other LLMs were not significant (P > 0.05). (Supplementary Materials 5)

## Discussion

To the best of our knowledge, this is the first study to evaluate the performance of the four most advanced LLMs—GPT-4o, GPT-4, Gemini 1.5 Pro, and Claude 3 Opus—on the JNME.

All four LLMs that we tested passed the JNME 2018. GPT-4o, GPT-4, and Gemini 1.5 Pro passed the 2024 JNME, whereas GPT-4 did not. In terms of the overall accuracy for both the 2018 and 2024 examinations, GPT-4o achieved an accuracy of 89.2%, significantly outperforming GPT-4 (76.8%), Gemini 1.5 Pro (80.1%), and Claude 3 Opus (82%) by approximately 10%. Contrary to our hypothesis, GPT-4o, GPT-4, and Gemini 1.5 Pro exhibited higher accuracy in the 2024 exam than in the 2018 exam, although the differences were not statistically significant. This suggests that the performance of the LLMs was not affected by the cutoff date. This can be attributed to two factors. First, the training data for LLMs were primarily in English, with limited Japanese data ^34^. This finding indirectly supports the reliability of the present study. Second, the 2018 exam had a higher proportion of difficult and normal questions (50.8%) than the 2024 exam (37.5%). We believe that the difference in question difficulty between the 2018 and 2024 examinations led to higher accuracy for LLMs in the 2024 exam.

Previous studies showed that GPT-4’s performance on image-based questions is inferior to that on non-image-based questions ^35, 36^. We investigated the performance of more advanced LLMs for image-based questions. Consistent with previous studies, we found that the advanced LLMs performed significantly worse on image-based questions than on non-image-based questions.

The performance gaps for GPT-4o, GPT-4, Gemini 1.5 Pro, and Claude 3 Opus were 11.8%, 12.7%, 7.4%, and 6.2%, respectively. Notably, GPT-4o achieved an accuracy rate of 92.2% for non-image-based questions, surpassing 90%. The other three LLMs also achieved accuracy rates greater than 80% for non-image-based questions. Conversely, the accuracy rates of the LLMs for image-based questions were all less than or equal to 80%. This indicates that, at the current stage, LLMs’ ability to recognize medical images is still suboptimal, posing a high risk of premature use in medical imaging analysis and diagnosis. Therefore, improving LLMs’ image recognition capabilities is essential for their future development.

We categorized all questions based on human medical students’ accuracy into three categories: easy, normal, and difficult, and tested the LLMs accordingly. All four LLMs performed significantly better on easy questions than on normal questions and significantly better on normal questions than on difficult questions, with performance gaps of approximately 10%. This indicates that similar to previous LLMs, the performance of advanced LLMs is significantly affected by the difficulty level of questions ^19, 37–42^. Notably, GPT-4o achieved an accuracy rate of 95.0% (421/443) for easy questions and had the highest score for any category in this study. This also represents the highest performance observed in large-scale (over 100 questions) medical exam testing studies of LLMs to date, surpassing the 95% passing threshold considered adequate for use as a knowledge resource in medical education ^11, 22^. We also calculated the accuracy of GPT-4o for easy, non-image-related questions, which was 96.2% (329/342). This suggests that for some fundamental medical questions, medical students can use the GPT-4o as an effective knowledge resource.

Typically, the performance of LLMs on general questions reflects their accuracy as knowledge sources in medical education, whereas their performance on clinical questions reflects their capability in clinical diagnosis. By comparing the performance of the LLMs on general and clinical questions, we found that none of the four LLMs exhibited significant differences. This suggests that whether a question is general or clinical is not a potential factor influencing LLM performance. Consequently, LLMs have not demonstrated a significant advantage in medical education or clinical diagnosis.

A previous study reported the performance of GPT-4 and GPT-3.5, across different medical specialties, but did not specify which specialties the LLMs performed better or worse than their overall performance ^43^, while our study fills this gap. By comparing the performance differences in the LLMs across different medical specialties, we found that the accuracy rates of the LLMs in Gastroenterology and Hepatology (P<0.001), Pulmonology (P=0.046), and Hematology (P=0.02) were significantly lower than the average accuracy. Notably, Gastroenterology and Hepatology ranked last for GPT-4o, GPT-4, and Claude 3 Opus, and second to last for Gemini 1.5 Pro. By contrast, the accuracy rates for Cardiology (P<0.001), Emergency Medicine (including Toxicology), Anesthesiology, and Intensive Care (P<0.01), and Immunology and Rheumatology (P=0.03) were significantly higher than the average accuracy. These results suggest that LLMs are not suitable for use in medical education or clinical diagnosis in Gastroenterology and Hepatology, Pulmonology, and Hematology. To explore the reasons for the performance differences in LLMs across specialties, we analyzed the correlation between the accuracy rates in each specialty and the number of WOS articles. The results show a positive correlation between LLM performance and the number of articles in each specialty, with Gemini 1.5 Pro showing the strongest correlation. This indicates that, in the medical field, the number of articles is one of the primary sources of LLM learning, and LLMs are more likely to perform poorly in areas with insufficient articles.

### Limitation

Previous studies have employed various prompts to test LLMs. However, there is currently no standardized prompt template recognized by researchers or LLM developers to enhance LLM performance. In addition, to simulate the real-world use of LLMs by users, our study did not use prompts and only tested the performance of LLMs in their default settings. However, a previous study suggested that prompts could improve the LLM performance to a certain extent ^22^. We hypothesized that the use of prompts would bring LLMs, particularly GPT-4, closer to the 95% threshold. We plan to test this hypothesis in future studies.

Furthermore, in analyzing the performance of LLMs across different medical specialties, we did not use specific exams for each specialty, but instead categorized questions from comprehensive medical licensing exams by specialty. This resulted in fewer question samples from certain specialties. A more detailed assessment of the LLM performance in each medical specialty requires testing with a specific examination of each medical specialty. However, considering that Fisher’s exact test evaluates significance based on P-values, which are a factor in sample size, we believe that the results of our study indicate that significantly higher or lower than average accuracy in certain specialties are reliable.

Finally, previous study found that GPT-4V, even when selecting the correct option for image-based MCQs, may provide incorrect reasoning ^44^. However, GPT-4V is not one of the tested LLMs in this study. We hypothesize that more advanced LLMs will address this issue. We will verify this hypothesis in future study.

## Conclusion

This study evaluated the performances of GPT-4o, GPT-4, Gemini 1.5 Pro, and Claude 3 Opus in JNME. Regardless of the overall accuracy or accuracy across various categories, GPT-4o demonstrated significantly higher accuracy than the other three LLMs. GPT-4o achieved an impressive overall accuracy of 89.2%, and in the category of easy questions, it reached a threshold of 95%, which can be considered an effective knowledge resource. This marks a milestone in the development of LLMs. All four LLMs showed significantly higher accuracies for non-image-based questions than for image-based questions. Among them, GPT-4o achieved a non-image-based question accuracy of 92.2%, which is close to the 95% passing threshold. For more advanced LLMs, question difficulty remains a major factor affecting accuracy. All four LLMs performed poorly in the medical specialty of Gastroenterology and Hepatology. General and clinical questions did not affect the LLM performance. Finally, the four LLMs showed a positive correlation between the number of publications and the performance in different medical specialties, with Gemini 1.5 Pro showing the strongest correlation.

## Supporting information

Supplementary Materials 1 in the manuscript.

Supplementary Materials 2 in the manuscript.

Supplementary Materials 3 in the manuscript.

Supplementary Materials 4 in the manuscript.

Supplementary Materials 5 in the manuscript.

## Author Contributions

Study concepts/study design, M.L., T.O., T.K.; manuscript drafting or manuscript revision for important intellectual content, all authors; approval of the final version of the submitted manuscript, all authors; agrees to ensure any questions related to the work are appropriately resolved, all authors; literature research, M.L., H.O.; experimental studies, M.L., Z.D., W.H., E,F.; JNME questions preparing/advices, E.F., Z.D.; data interpretation and statistical analysis, M.L., W.H., H.O.; and manuscript editing, all authors.

## Data Availability

All data produced in the present work are contained in the manuscript.

## Acknowledgments

None.

## Funding/Support

This work was supported by JSPS KAKENHI Grant Number 24KJ0830.

### Other disclosures

None

### Ethical approval

Not applicable

### Disclaimers

None

### Previous presentations

None

---

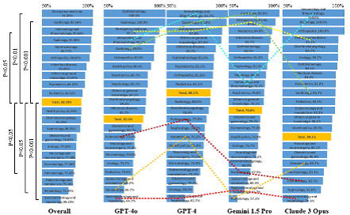

