## Supplementary Materials 1 in the manuscript. for "Performance of Advanced Large Language Models (GPT-4o, GPT-4, Gemini 1.5 Pro, Claude 3 Opus) on Japanese Medical Licensing Examination: A Comparative Study"

**Supplementary Materials 1. Characteristic of JNME questions.**

| Categories | | | Number |
| --- | --- | --- | --- |
| Years | **JNME 2018** | | **390** |
|  | **JNME 2024** | | **400** |
|  | **Total** | | **790** |
| General or clinical questions | **General questions** | **JNME 2018** | **146** |
|  |  | **JNME 2024** | **147** |
|  |  | **Total** | **293** |
|  | **Clinical questions** | **JNME 2018** | **253** |
|  |  | **JNME 2024** | **244** |
|  |  | **Total** | **497** |
| With or without images | **Questions with image** | **JNME 2018** | **107** |
|  |  | **JNME 2024** | **92** |
|  |  | **Total** | **199** |
|  | **Questions without image** | **JNME 2018** | **283** |
|  |  | **JNME 2024** | **308** |
|  |  | **Total** | **591** |
| MCQs or calculation questions | **MCQs** | | **784** |
|  | **Calculation questions** | | **6** |
| Level of difficulty | **Easy** | **JNME 2018** | **192** |
|  |  | **JNME 2024** | **251** |
|  |  | **Total** | **433** |
|  | **Normal** | **JNME 2018** | **122** |
|  |  | **JNME 2024** | **95** |
|  |  | **Total** | **217** |
|  | **Difficult** | **JNME 2018** | **76** |
|  |  | **JNME 2024** | **54** |
|  |  | **Total** | **130** |
| Medical specialty | **Others or general knowledge** | | **142** |
|  | **Health policy, epidemiology, and public health** | | **69** |
|  | **Obstetrics and gynecology** | | **57** |
|  | **Cardiology** | | **55** |
|  | **Infectious disease** | | **43** |
|  | **Psychiatrics** | | **28** |
|  | **Endocrinology and metabolism** | | **26** |
|  | **Pediatrics** | | **53** |
|  | **Emergency medicine (including toxicology), anesthesiology and intensive care** | | **41** |
|  | **Immunology and Rheumatology** | | **24** |
|  | **Gastroenterology and hepatology** | | **51** |
|  | **Nephrology** | | **21** |
|  | **Hematology** | | **21** |
|  | **Otorhinolaryngology** | | **19** |
|  | **Pulmonology** | | **40** |
|  | **Ophthalmology** | | **17** |
|  | **Radiology** | | **5** |
|  | **Neurology and neurosurgery** | | **36** |
|  | **Orthopedics** | | **11** |
|  | **Dermatology** | | **12** |
|  | **Urology** | | **19** |
