## Supplementary Materials 4 in the manuscript. for "Performance of Advanced Large Language Models (GPT-4o, GPT-4, Gemini 1.5 Pro, Claude 3 Opus) on Japanese Medical Licensing Examination: A Comparative Study"

**Supplementary Materials 4. Accuracy rate of each LLM on different medical specialty.**

|  | GPT-4o | GPT-4 | Gemini 1.5 PRO | Claude 3 Opus | Average |
| --- | --- | --- | --- | --- | --- |
|  | Correct number, rate | Correct number, rate | Correct number, rate | Correct number, rate | Correct rate |
| Total questions (n=790) | 89.2% | 76.8% | 80.1% | 82.0% | 82.0% |
| Others or general knowledge (n=142) | 128, 90.1% | 111, 78.2% | 124, 87.3% | 120, 84.5% | 85.0% |
| Health policy, epidemiology, and public health (n=69) | 62, 89.9% | 51, 73.9% | 52, 75.4% | 61, 88.4% | 81.9% |
| Obstetrics and gynecology (n=57) | 49, 86% | 43, 75.4% | 40, 70.2% | 46, 80.7% | 78.1% |
| Cardiology (n=55) | 51, 92.7% | 50, 90.9% | 50, 90.9% | 50, 90.9% | 91.4% |
| Infectious disease (n=43) | 40, 93% | 36, 83.7% | 36. 83.7% | 36, 83.7% | 86.0% |
| Psychiatry (n=28) | 26, 92.9% | 22, 78.6% | 23, 82.1% | 24, 85.7% | 84.8% |
| Endocrinology and metabolism (n=26) | 24, 92.3% | 17, 65.4% | 19, 73.1% | 18, 69.2% | 75.0% |
| Pediatrics (n=53) | 49, 92.5% | 45, 84.9% | 43, 81.1% | 39, 73.6% | 83.0% |
| Emergency medicine (including toxicology), anesthesiology and intensive care (n=41) | 39, 95.1% | 36, 87.8% | 36, 87.8% | 39, 95.1% | 91.5% |
| Immunology and Rheumatology (n=24) | 24, 100% | 19, 79.2% | 22, 91.7% | 22, 91.7% | 90.7% |
| Gastroenterology and hepatology (n=51) | 36, 70.6% | 32, 62.7% | 32, 62.7% | 33, 64.7% | 65.2% |
| Nephrology (n=21) | 17, 81% | 17, 81% | 16, 76.2% | 18, 85.7% | 81.0% |
| Hematology (n=21) | 18, 85.7% | 12, 57.1% | 16, 76.2% | 14, 66.7% | 71.4% |
| Otorhinolaryngology (n=19) | 18, 94.7% | 13, 68.4% | 15, 78.9% | 16, 84.2% | 81.6% |
| Pulmonology (n=40) | 33, 82.5% | 27, 67.5% | 31, 77.5% | 30, 75% | 75.6% |
| Ophthalmology (n=17) | 16, 94.1% | 14, 82.4% | 14, 82.4% | 17, 100% | 89.7% |
| Radiology (n=5) | 5, 100% | 4, 80% | 4, 80% | 5, 100% | 90.0% |
| Neurology and neurosurgery (n=36) | 31, 86.1% | 26, 72.2% | 27, 75% | 27, 75% | 77.1% |
| Orthopedics (n=11) | 11, 100% | 9, 81.8% | 9, 81.8% | 10, 90.9% | 88.6% |
| Dermatology (n=12) | 10, 83.3% | 9, 75% | 9, 75% | 9, 75% | 77.1% |
| Urology (n=19) | 18, 94.7% | 14, 73.7% | 13, 68.4% | 14, 73.7% | 77.6% |
