## Supplementary Materials 5 in the manuscript. for "Performance of Advanced Large Language Models (GPT-4o, GPT-4, Gemini 1.5 Pro, Claude 3 Opus) on Japanese Medical Licensing Examination: A Comparative Study"

**Supplementary Materials 5. Correlation between accuracy and the number of publications of each medical special.**

|  | GPT-4o | | | GPT-4 | | | Gemini 1.5 PRO | | | Claude 3 Opus | | |
| --- | --- | --- | --- | --- | --- | --- | --- | --- | --- | --- | --- | --- |
| Document categories | All-type documents | Articles | Open access articles | All-type documents | Articles | Open access articles | All-type documents | Articles | Open access articles | All-type documents | Articles | Open access articles |
| Cor | 0.11 | 0.29 | 0.22 | 0.19 | 0.29 | 0.21 | 0.42 | 0.47 | 0.44 | 0.21 | 0.35 | 0.27 |
| P-value (95%CI) | 0.61 (-0.34-0.53) | 0.21 (-0.17-0.65) | 0.34 (-0.24-0.6) | 0.42 (-0.27-0.58) | 0.22 (-0.17-0.64) | 0.37 (-0.26-0.6) | 0.07 (-0.03-0.73) | 0.03 (0.04-0.76) | 0.05 (0-0.74) | 0.37 (-0.25-0.6) | 0.13 (-0.11-0.68) | 0.24 (-0.19-0.64) |
